# A Rapid Assessment of Stakeholder Agreement with Brazil’s New Gender-Affirming Care Restrictions

**DOI:** 10.1101/2025.09.17.25335994

**Authors:** Bruna Caruso Mazzolani, Igor Longobardi, Alexandre Saadeh, Hamilton Roschel, Bruno Gualano

## Abstract

Transgender youth and their caregivers are rarely included in regulatory processes concerning gender-affirming care, despite being directly affected. In April 2025, the Brazilian Federal Council of Medicine (CFM) issued Resolution 2.427/2025, which banned the use of puberty blockers in all minors, prohibited hormone therapy before age 18, and introduced stricter age thresholds for surgical interventions. This study aimed to assess stakeholder perspectives regarding these new restrictions. We conducted a cross-sectional survey between May and June 2025 with patients and caregivers from Brazil’s largest outpatient gender identity service. A total of 54 transgender youth (mean age 13.6 ± 2.0 years) and 116 caregivers completed the survey independently. Participants rated their level of agreement with each provision of the resolution using a 6-point Likert scale. Findings demonstrated overwhelming opposition to the new regulations. Among youth, 92.6% strongly disagreed with the ban on puberty blockers, 94.4% opposed restrictions on hormone therapy before 18, and 59.3% disagreed with surgical age limits. Caregivers reported similar opposition, with 93.1% rejecting the ban on puberty blockers, 89.7% rejecting restrictions on hormone therapy, and 61.2% disagreeing with surgical restrictions. Importantly, opposition was consistent across all stages of treatment engagement, including among those who had not initiated or had declined medical interventions, suggesting that disagreement is not solely attributable to personal investment in treatment. These findings highlight a substantial misalignment between regulatory policy and the perspectives of directly affected youth and families. Incorporating patient and caregiver voices, alongside clinical evidence, is essential to ensure that policies remain ethically sound and responsive to stakeholder needs.

**Public significance statement:** This study found that over 90% of transgender youth and their caregivers in Brazil strongly oppose new bans on gender-affirming medical care, revealing a significant disconnect between regulatory policies and the lived experiences of those most affected. These findings underscore the critical importance of incorporating patient and family perspectives into healthcare policy to ensure it is both ethical and effective.

## Introduction

Globally, regulatory frameworks governing gender-affirming medical care for transgender and gender-diverse adolescents are undergoing significant revision, often toward more restrictive policies (Longobardi et al., 2025). In the US, over 20 states have enacted laws banning access to puberty blockers and hormone therapy for minors (House, 2025). In the UK, the National Health Service now limits puberty suppression to research settings only (Cass, 2024; NHSE, 2024). A frequently cited rationale for these policy shifts is the perceived lack of high-certainty evidence regarding long-term outcomes and the capacity for informed consent among adolescents.

In April 2025, Brazil’s Federal Council of Medicine (CFM) promulgated Resolution No. 2.427/2025, introducing substantial changes to the provision of care for transgender youth (CFM, 2025). This resolution prohibits the use of puberty blockers for all children and adolescents, bans hormone therapy for individuals under the age of 18, and imposes age restrictions on surgical interventions. Furthermore, it mandates that transgender individuals receive care from medical specialists associated with their sex assigned at birth. This new regulatory stance represents a departure from previous clinical protocols followed in Brazil, which were consonant with international guidelines from organizations such as the World Professional Association for Transgender Health (WPATH) and the Endocrine Society (Coleman et al., 2022; Hembree et al., 2017; CFM, 2019) (more details in Methods).

While the call for more rigorous long-term data is a recognized priority within the field, the implementation of restrictive policies based on evidence limitations itself raises questions. A key concern is whether such sweeping regulatory actions adequately consider the potential harms of delaying or denying care, as well as the perspectives of those directly affected. Eliciting stakeholder views is considered a critical component of health services research and ethical policy formation, ensuring that regulations are informed not only by evolving evidence but also by patient values and lived experience. This study, therefore, aimed to conduct a cross-sectional survey to assess the level

of agreement with the provisions of CFM Resolution 2.427/2025 among transgender youth and caregivers receiving care at a major Brazilian gender identity service.

## Methods

This cross-sectional study was conducted between May and June 2025 and was approved by the local ethics committee (Commission for Analysis of Research Projects, CAPPesq; approval: 83553224.2.0000.0068). Participants included transgender youth aged 10–18 years and their caregivers, as well as caregivers of transgender children under 10, all receiving care at the Transdisciplinary Gender Identity and Sexual Orientation Service at the Clinical Hospital of the University of São Paulo Medical School. Recruitment occurred in person during clinical visits or remotely via phone, messaging apps and email with prior authorization.

All youth (n=153) and caregivers (n=239) from our outpatient service were invited to participate. They were instructed to complete the survey independently to ensure individual responses. Written informed consent was obtained from caregivers and assent from participating youth.

Before the resolution 2.427/2025, gender-affirming care for minors in Brazil followed internationally recognized clinical protocols (Coleman et al., 2022; Hembree et al., 2017; CFM, 2019). Puberty blockers (GnRH analogs) were offered from Tanner stage ≥2 after comprehensive biopsychosocial evaluation by a multidisciplinary team. Eligible patients started hormone therapy (estrogen or testosterone) typically around age 16, with informed consent or assent and parental permission. Care was aligned with guidelines from the WPATH and the Endocrine Society (Coleman et al., 2022; Hembree et al., 2017; CFM, 2019), and provided exclusively through the public health system (SUS) in federally accredited university hospitals with specialized teams. The new resolution introduced the following key changes: (1) Prohibition of puberty blockers for all children and adolescents; (2) Ban on hormone therapy before age 18; (3) Restriction of gender-affirming surgeries to individuals 18 or older; sterilizing procedures permitted only from age 21; and (4) Requirement that transgender individuals with reproductive anatomy associated with their sex assigned at birth (e.g., uterus, ovaries, penis, testicles) remain under care of specialists aligned with that sex. Participants completed an online survey in which they rated their level of agreement with the provisions of the new resolution using a 6-point Likert scale. All participants were fully informed about the resolution’s implications and understood that care at the clinic would continue to be provided in accordance with the new regulations. Sociodemographic and clinical data, including mental health conditions, were obtained from medical records. Data are reported descriptively.

## Results

A total of 54 youth and 116 caregivers of children and adolescents completed the survey. Among the 116 caregiver respondents, 48 were caregivers of youth who also completed the survey, whereas 55 were caregivers of children or adolescents who did not respond the survey, and 13 were secondary caregivers (i.e., more than one caregiver per child or adolescent). Youth and caregiver responses were analyzed independently to capture individual perspectives, acknowledging partial overlap between the groups.

Youth participants had a mean age of 13.6 ± 2.0 years, most identified as male (36/54, 68.5%), 23 (42.6%) were undergoing puberty suppression, and 4 (7.4%) were receiving hormone therapy. Among the transgender children and adolescents represented by caregivers (n=103), the mean age was 11.8 ± 2.7 years, 67 (65.1%) were identified as male, 32 (31.1%) were under puberty suppression, and 2 (1.9%) were receiving hormone therapy (Table 1). Most caregiver respondents identified as female (92/116, 79.3%).

**Table 1.**
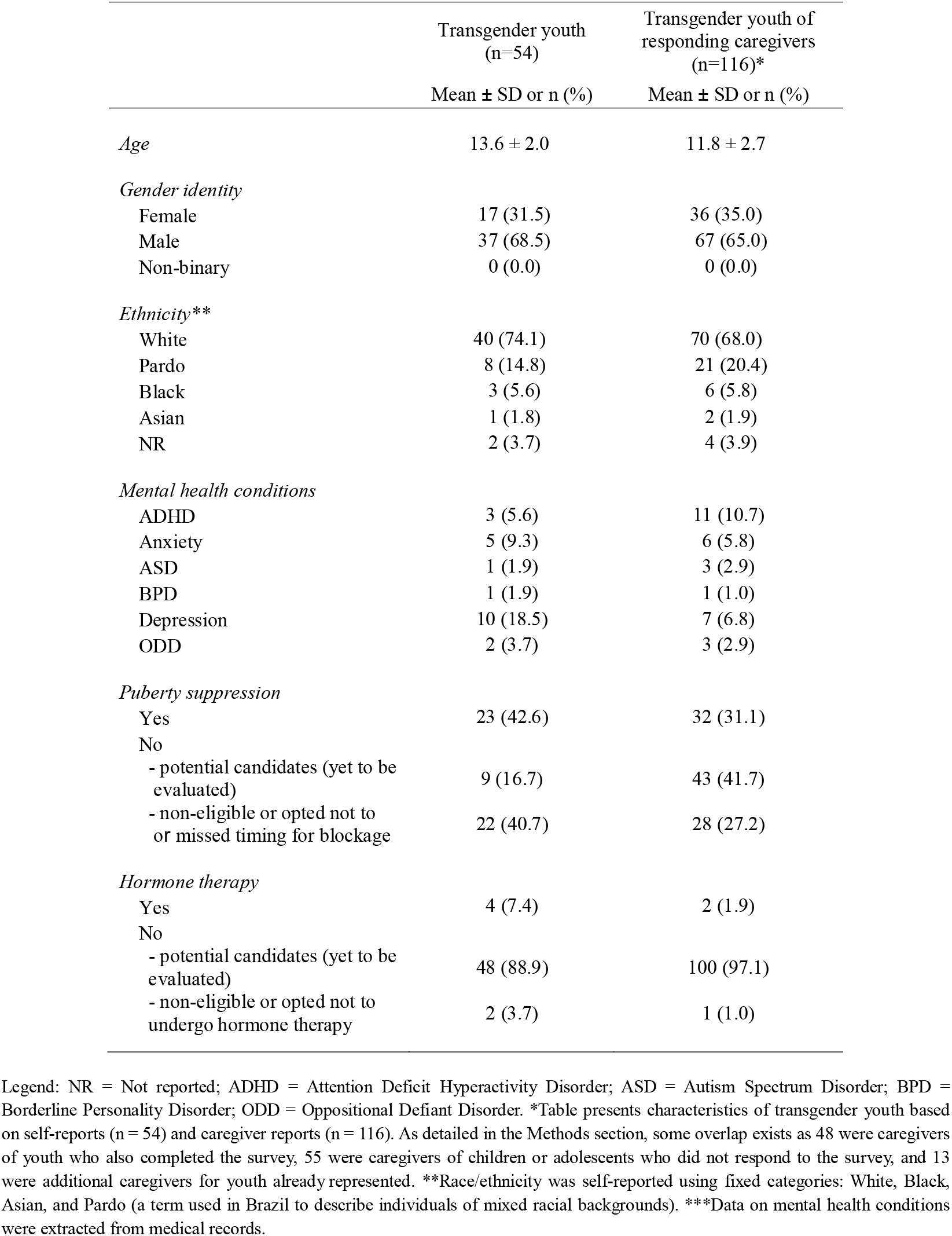
Demographic and Clinical Characteristics of Transgender Youth (n=54) and Transgender Youth Whose Caregivers Responded to the Survey (n=116).

Fifty out of 54 transgender youth (92.6%) strongly disagreed with the ban on puberty blockers, while 51 (94.4%) strongly disagreed with the restriction on hormone therapy, 32 (59.3 %) strongly disagreed with the age restrictions on gender-affirming and sterilizing surgeries and 18 (33.3%) opposed the requirement to receive care from specialists based on their sex assigned at birth (Table 2).

**Table 2.** Transgender Youth (n=54) and Caregivers (n=116) agreement with the new norms introduced by the resolution.

| Population | Provision summary | 0 | 1 | 2 | 3 | 4 | 5 |
| --- | --- | --- | --- | --- | --- | --- | --- |
| Transgender Youth<br>n (%) | Puberty blockers prohibition <sup>1</sup> | 50 (92.6) | 2 (3.7) | 0 (0.0) | 0 (0.0) | 0 (0.0) | 2 (3.7) |
|  | Hormone therapy restrictions <sup>2</sup> | 51 (94.4) | 1 (1.9) | 0 (0.0) | 0 (0.0) | 0 (0.0) | 2 (3.7) |
|  | Surgical age restrictions <sup>3</sup> | 32 (59.3) | 4 (7.4) | 4 (7.4) | 7 (13.0) | 3 (5.6) | 4 (7.4) |
|  | Specialist care requirements<br>(based on sex assigned at birth) <sup>4</sup> | 9 (16.7) | 0 (0.0) | 9 (16.7) | 15 (27.8) | 5 (9.3) | 16 (29.6) |
| Caregiver<br>n (%) | Puberty blockers prohibition | 108 (93.1) | 5 (4.3) | 0 (0.0) | 0 (0.0) | 2 (1.7) | 1 (0.9) |
|  | Hormone therapy restrictions | 104 (89.7) | 5 (4.3) | 6 (5.2) | 0 (0.0) | 0 (0.0) | 1 (0.9) |
|  | Surgical age restrictions | 71 (61.2) | 7 (6.0) | 10 (8.6) | 8 (6.9) | 4 (3.5) | 16 (13.8) |
|  | Specialist care requirements<br>(based on sex assigned at birth) | 36 (31.0) | 6 (5.2) | 16 (13.8) | 9 (7.8) | 8 (6.9) | 41 (35.3) |

Legend: 6-point Linkert scale (0 means strongly disagree and 5 means strongly agree)
<sup>1</sup> Prescription prohibited for transgender/gender-diverse children and adolescents;
<sup>2</sup> Use prohibited before age 18 to induce gender-aligned physical characteristics;
<sup>3</sup> All gender-affirming surgeries: ≥18 years; Surgeries causing infertility: ≥21 years;
<sup>4</sup> Transgender men with uterus/ovaries: Gynecologist/Transgender women with penis/testicles: Urologist.

Most caregivers strongly disagreed with the prohibition of puberty blockers (108/116 [93.1%]), the restriction on hormone therapy for minors (104/116 [89.7%]), and the restriction of gender-affirming surgeries before age 18 and sterilizing procedures before age 21 (71/116 [61.2%]). Half of the caregivers disagreed (58/116 [50.0%]) with the mandate that transgender individuals should be treated by specialists of their sex assigned at birth (Table 2).

## Discussion

In this survey, most caregivers and transgender youth disagreed with recent restrictions imposed by the CFM on gender-affirming care for minors. The high levels of disagreement, especially with the blanket ban on puberty blockers and the deferral of hormone therapy until age 18, suggest a significant divergence between the perspectives of those directly affected and the assumptions underlying the new regulatory framework.

These findings are particularly relevant in the context of similar regulatory trends adopted in the US and UK, where access to gender-affirming care for adolescents has also been restricted (House, 2025; NHSE, 2024). Existing evidence from prospective and observational studies suggests that gender-affirming interventions, when provided under appropriate clinical oversight, are associated with reductions in depression, anxiety, and suicidality, and with low rates of regret or detransition (Costa et al., 2015; de Vries et al., 2014; Olson et al., 2024; Tordoff et al., 2022; van der Loos et al., 2022), although some reports (e.g., Cass Review (Cass, 2024)) have indicated the low quality or limited certainty of the evidence for such interventions in minors. While calls for more long-term data are valid, capturing the voices of directly affected stakeholders is highly relevant, as studies from the U.S. similarly show that care bans provoke fear and distress among families (Abreu et al., 2022; Kidd et al., 2025). Such perspectives are a key component of evidence-based medicine, which emphasizes the integration of research evidence, clinical expertise, and patient values. Framing our findings in this context highlights the need for regulatory debates to consider stakeholder voices alongside ongoing evaluations of the clinical evidence base.

In this sense, a strength of this study is its timely assessment of stakeholder perspectives in the immediate aftermath of a major policy change, collecting reactions from both caregivers and patients actively engaged in gender-affirming care at the largest clinic in Brazil. The generalizability of these findings is constrained by the recruitment of participants from a single, specialized clinic. As a result, the perspectives captured may disproportionately reflect individuals and families already engaged with, and committed to, the established model of gender-affirming care. Future research would benefit from incorporating a broader range of perspectives, including those of transgender individuals who have not sought medical care, those who have discontinued treatment, and families who have been hesitant about or declined care. This limitation, however, must be interpreted in light of the structure of the Brazilian public health system, in which such care is exclusively centralized within federally accredited, university-based clinics. Within this context, the present cohort is representative of the national patient population that actively accesses this standardized care pathway. Importantly, the consistent opposition observed across all stages of treatment engagement, including among individuals who had not initiated medical interventions or had decided against pursuing blockade, suggests that disagreement with the resolution cannot be attributed solely to personal investment in ongoing treatment. While broader perspectives remain essential for comprehensive policy discussions, the present data provide a critical and informed account from primary stakeholders directly affected by the regulatory change.

In conclusion, our results demonstrate overwhelming opposition to recent regulatory changes among directly affected youth and their caregivers, highlighting a critical misalignment between policy and stakeholder perspectives. Regulatory decisions in transgender health care may benefit from being informed not only by the best available clinical evidence but also by the lived experiences and values of patients and caregivers, consistent with the principles of evidence-based medicine. Such integration can help ensure that policies remain clinically grounded while also responsive to the needs of those most directly affected.

## Data Availability

The data that support the findings of this study are available from the corresponding author upon reasonable request.

## Acknowledgements

The authors are supported by the São Paulo Research Foundation (FAPESP), the National Council for Scientific and Technological Development (CNPq), and the Coordination for the Improvement of Higher Education Personnel (CAPES). Funders had no role in the preparation of this manuscript.

